# Effects of home environmental, behavioural and domestic activities on the risk of home injuries in French adults: Results from a prospective study

**DOI:** 10.1101/2022.07.18.22277761

**Authors:** Madelyn Yiseth Rojas Castro, Marta Avalos, Benjamin Contrand, Marion Dupuy, Catherine Sztal-Kutas, Ludivine Orriols, Emmanuel Lagarde

## Abstract

Prospective home injuries (HIs) and detailed exposure information are generally scarce to study risk factors. We assessed the associations between physical household environment, behaviours and Do-It-Yourself (DIY), gardening and domestic activities and HI’s risk in a prospective cohort study. The MAVIE observatory is an e-cohort conducted among volunteers of the French general population. Poisson mixed models were fitted and Risk Ratios estimated to assess the determinant of the number of HIs prospectively recorded during the follow-up. A total of 6146 dwelling adults aged 15 or more were followed up for 4.0 years on average and 12% reported at least one HI during follow-up. Adjusting on socio-demographics variables and self-perception of physical and mental health, no characteristic of the physical environment measured at baseline were associated with HI risk. Storing household products out of their original packaging, DIY activities and using a stool to reach high places were significantly associated with HI with attributable fractions of 12.1%, 6.4% and 6.9% respectively.

## Introduction

Home is the environment in which most unintentional injuries occur (Keall et al., 2011; Runyan et al., 2005). Home injuries (HIs) represent a substantial health burden worldwide, approximately one-third of the global burden of injuries (Haagsma et al., 2016; Keall et al., 2011; WHO, 2018). In the EU-28, HIs caused an annual average of 85 thousand fatalities, 1.5 million hospital admissions and 9 million emergencies between 2012 and 2014 (Eurosafe, 2016). In the United States, 75% of all preventable injury-related deaths occurred in homes and communities in 2019 (National Safety Council, 2019). This large HI burden and the current interest in the effect of housing conditions on health disparities stresses the need to understand the effect of exposure to home environmental hazards, together with individual vulnerabilities and social determinants, on the risk of experiencing HI (Gielen et al., 2015; Keall et al., 2011; Swope & Hernández, 2019; WHO, 2018). Particular attention was paid to environmental barriers related to functional limitations in community-dwelling older adults (Ekström et al., 2016; Granbom et al., 2016; Tsuchiya et al., 2018).

Several studies have been conducted to identify injury hazards at home (Camilloni et al., 2011; Ferrante et al., 2013; Gielen et al., 2015; Keall et al., 2011) and to measure the effectiveness of hazard modification interventions and prevention strategies for the leading causes of home injury (i.e., home fires and burns, poisonings and falls) (Camilloni et al., 2011; Dionyssiotis, 2012; Farchi et al., 2006; Gillespie et al., 2003; Lord et al., 2006; Schaudt et al., 2019). Literature reviews showed strong evidence for an association between hazards at home and injuries (Carter et al., 1997; Keall et al., 2008; WHO, 2018). The certainty of the evidence is moderate to high for reducing hazards at home and moderate for the efficacy of hazard modification programs (WHO, 2018).

According to these studies, some confirmed home hazards related to injury risk are the absence of railings on stairways (Lord et al., 2006), the height of rails and the spaces between the stair rails (Keskinoglu et al., 2008), insufficient lighting (Camilloni et al., 2011; Farchi et al., 2006; Gillespie et al., 2003), deteriorated stairs (Gielen et al., 2015), slippery or uneven floors (Lord et al., 2006) (such as vinyl or linoleum (Talbot et al., 2005)), steps or uneven surfaces (Talbot et al., 2005), loose rugs or carpets (Gillespie et al., 2003; Lord et al., 2006), loose electrical wires on the bedroom floor {Healey1994}, unstable furniture and obstructed walkways (Gillespie et al., 2003; Lord et al., 2006), lack of window guards at the second level or higher (Keall et al., 2011), tap water that is too hot (Gielen et al., 2015), lack of working smoke alarms (Gielen et al., 2015; Keall et al., 2011), electrical hazards and sources of carbon monoxide (CO) (Gielen et al., 2015), having a garden (Ferrante et al., 2014) and having a balcony (Ferrante et al., 2014).

In the same sense, the evidence of the effectiveness of home hazard modifications on falls in the elderly is uncertain (Carter et al., 1997; Clemson et al., 1996; Gillespie et al., 2003, 2009, 2012; Lyons et al., 2003; Turner et al., 2017). Effectiveness varies, depending on interacting health factors (history of falls, limited mobility, poor balance, cognitive impairment) (Ambrose et al., 2013; Clemson et al., 1996; Lord et al., 2006), risk-taking factors, and the intervention methods used. For instance, grab rails/bars and slips/trips in the bathroom were described to adequately reduce falls in older people with reduced physical abilities (Gillespie et al., 2009, 2012).

Unsafe behaviours and at-risk behaviours in the domestic environment have been less studied. Participation in activities such as do-it-yourself (DIY), gardening and domestic activities have been associated with a higher incidence of HI in several original studies conducted in the general population (Ashby et al., 2007; Schaudt et al., 2019; Yiengprugsawan et al., 2012). Individual, behavioural and environmental factors influence higher risk levels. Knowledge of the relationships between home hazards and injury occurrence is still limited, principally due to insufficient data on exposure and potentially confounding factors (Gielen et al., 2015; Keall et al., 2011; WHO, 2018). Further research is required to identify new risk factors and understand the effect of environmental and behavioural factors, particularly among the most vulnerable and in other groups where studies are scarce (Gielen et al., 2015; Peeters et al., 2019).

The purpose of this study was to assess associations of a wide range of home environmental, behavioural and domestic activities factors on the risk of non-fatal HI in adults of all ages in the MAVIE cohort. The availability of prospective cohort-based data and the access to relevant exposure information (e.g., socioeconomic information, time spent at home, health status, history of injuries) allows us to study the most relevant associations, some of them unexplored, which can be helpful in the design of future interventions.

## Materials and methods

### Study design

The MAVIE cohort is a web-based prospective cohort study conducted in France, with a longitudinal follow-up of HLIs (Rojas et al., 2021). All households in France and French overseas territories were eligible to participate. The recruitment process began in November 2014. The present study analysed the data collected up to December 31, 2019.

Cohort management was entirely online. Participants were recruited through an email invitation sent to their insurees by three mutual insurance companies, press releases, social media, posters, and flyers. A household reference member was in charge of completing a web-based questionnaire for the household. Consenting members of each household were asked to provide individual information. Caregivers were invited to represent and participate on behalf of one older person (to address the foreseeable difficulties with computer use).

The inclusion criteria for participation in the MAVIE cohort were: i) residing in France; ii) being able to answer the questionnaires in French; iii) having access to and being able to use the Internet (at least the reference member). The baseline sample was defined as the participants aged 15 or over who answered at least one question of the individual inclusion questionnaire. We excluded those who lived in hospitals and retirement or long-term care institutions. We only considered participants whose follow-up could be confirmed (by injury or non-injury report, questionnaire update, death or study leave request). We also excluded those participants who did not answer the daily schedule questions.

Events notification change in household composition and moves could be reported at any time. Also, every three months, household reference members received an email reminder to report any injury event to any consenting household members during the follow-up. If no injury had occurred, a link in the email allows reporting with a single click.

### Home Injuries

This study focused on unintentional home injuries or its premises. We excluded all events involving illnesses or medical symptoms. We excluded events that occurred before or on the same date as the date of consent and those for which information about the type of medical care or the circumstances of occurrence was not reported. Finally, we excluded injuries that occurred during sleeping time, considered unlikely (Kopjar & Wickizer, 1996).

Data included activity and location, mechanisms and type of medical care of the injury event. We defined HIs as severe when emergency care or hospital inpatient care was required.

### Individual characteristics

Gender, age, self-perceived physical and mental health, history of previous injury (excluding sports injuries) over the past 12 months, education status, household incomes, living situation, and alcohol consumption were studied at baseline.

Self-perceived mental health and self-perceived physical health were both self-rated using a visual analogue scale (1□=□*Poor health*, 10□=□*Excellent health*). The responses were classified as: excellent health (8 to 10), good health (5 to 7), and poor health (1 to 4). Occupational status groups were *unemployed, homemakers, and retirees* and *students and employees*. Responses to education status were assigned to ‘*educational attainment levels by age group*’. We considered a low level of education in high school or vocational studies, or lower for participants aged 20 to 54, and primary studies or lower for other ages. Annual household incomes were assigned to classes according to the French population’s percentiles reported in 2015 (low: ≤ 30^th^ percentile, middle: 30th-80th percentile, high: ≥ 80th percentile). The categories of the frequency of alcohol consumption were less than two times a week, and two or more times a week.

Participants were asked to report their ‘*typical daily schedule’* on a weekday and on a weekend day, the time spent at home and in different domestic settings (dining room, bedroom, garden, etc.). The average duration was also reported for domestic work, gardening, and do-it-yourself (DIY) activities. Time spent on each activity was categorized as *frequent* (over the 75th percentile), *occasional* or *null*.

#### Household characteristics and behaviours

Household level questions focused on the collection of data on general residence characteristics (location of the residence, type of housing, type of dwelling, age of construction, dwelling area), characteristics of the living environment, including habitat characteristics (interior and exterior), presence of domestic animals, safety equipment, heating system, electrical installations, physical hazards, other hazards and risky behaviours.

### Statistical analyses

Data were analysed using R version 3.6.1 (The R Foundation for Statistical Computing, Vienna, Austria)

We defined the follow-up period (FP) from the consent date to the date of the last information reported: injury or non-injury report, the questionnaire’s update date, the cohort exit date, the death date. We calculated the proportion of time spent awake at home (PTAH) as the proportion of hours on a weekday or weekend day when the person was neither away from home nor sleeping, for each of the declared households considering moves. We considered the *time-at-risk* as:

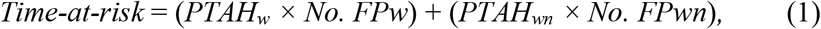

where *PTAH*_*w*_ was the time spent awake at home on a regular weekday, *PTAH*_*wn*_ on a regular weekend day, *No. FP*_*w*_ the number of weekdays in the follow-up period and *No. FP*_*wn*_ the number of weekend days. We calculated the proportion of *time-at-risk* as:

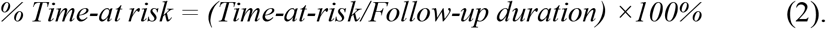

#### Risk factor analysis

Poisson mixed models were used to fit the number of HIs during the estimated exposure distribution in the person-time at risk for injury. Repeated measures are included for each individual to account for moves. Different follow-up times were assigned according to move dates. Nested random effects were included to consider the cluster structure of data in households. All models were fitted using the Template Model Builder R package (Brooks et al., 2017). Relative risks (RRs) were calculated conditionally upon the random effects, and the 95% confidence intervals were calculated using the profile method (Venzon & Moolgavkar, 1988). We adopted a complete case approach.

Confounding factors were *pre-selected* through ‘*Directed Acyclic Graphs’*. We fitted individual crude regression models to main exposures of interest and confounding factors. We conducted a variable selection procedure by steps and blocks of variables. In step 1, we adjusted each model by age, gender, history of previous injury, self-perceived physical and mental health status in all models, and we selected the most important associations (*P*_*adj*_ < 0.10). In step 2, we fitted the models to each of the exposures of interest, adjusting for significant adjustment variables selected in the previous step. In step 3, we fitted a fully adjusted model with the most important associations. Benjamini-Hochberg corrected *p-values* were computed (Benjamini & Hochberg, 1995). The same methodology was used to construct models stratified by two distinct age groups (15 to 49 years old and 50 years old and over). The attributable fraction (AF) was estimated using the adjusted RR of the fully adjusted models, as suggested by Flegal and colleagues, 2006 (Flegal et al., 2006).

Models were validated with appropriate diagnostic tools, ruling out problems of misspecification, such as over/under dispersion and zero inflation.

## Results

Between November 2014 and December 2019, 29931 potential participants were registered in the MAVIE Observatory by the reference members. 41% signed the informed consent and responded to baseline questionnaires. Among them, 11420 were 15 years old or more, 9429 were household reference members representing themselves and/or another household member. Sixty-six participants reported living in a retirement institution, and 3524 participants did not provide follow-up data. Another 1684 participants did not provide their *daily schedule* (Figure 1). The sample size was 6146 participants from 5122 households.

**Figure 1.**
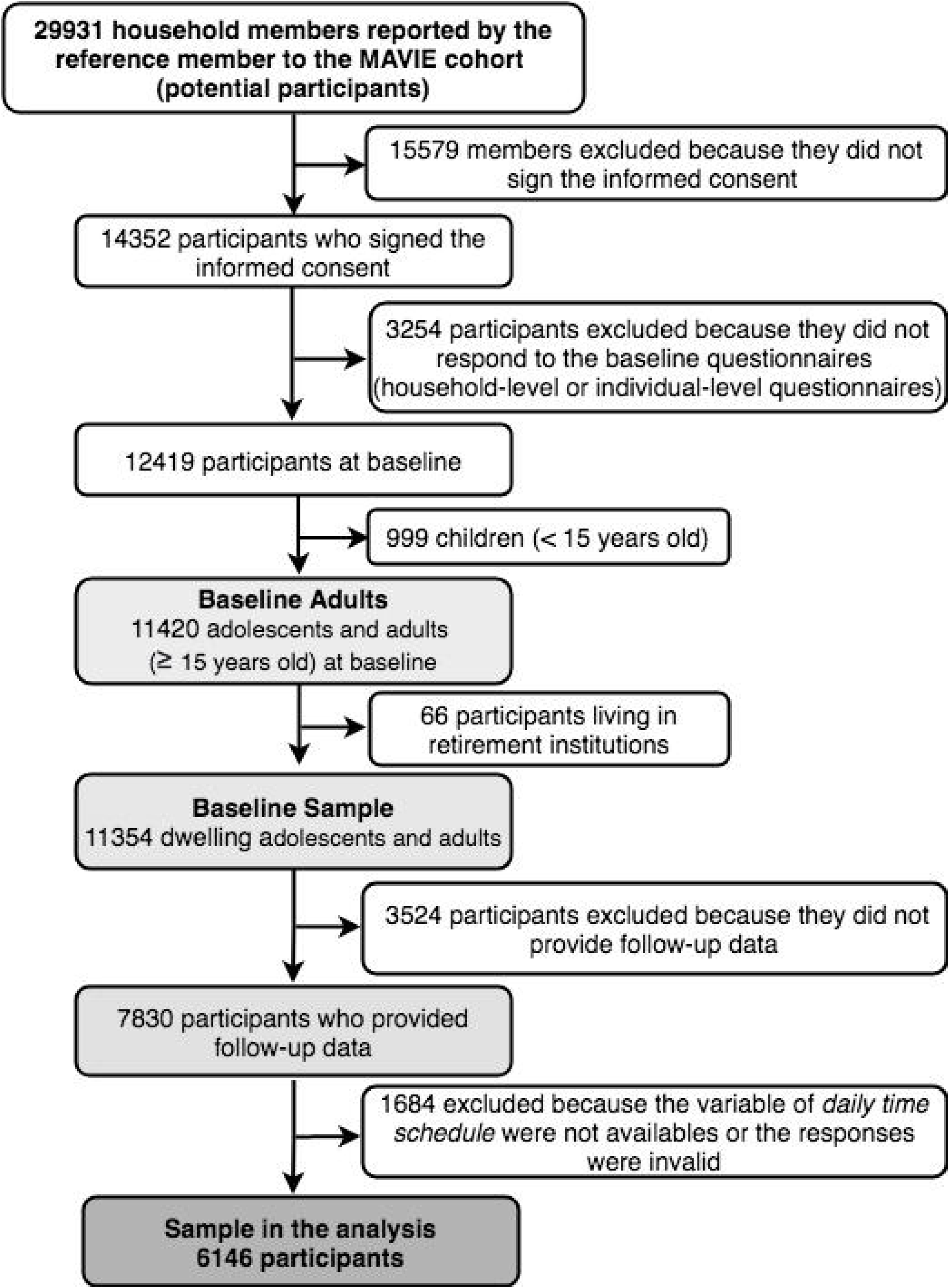
Flow diagram for the selection of study participants.

The median follow-up was 4.0 years (Q1=3.6, Q3=4.5) and the máximum 5.1 years. The overall follow-up was 24480 person-years. The estimated loss to follow-up was 13%.

Among the 6146 individuals of the study, 244 left the cohort during the follow-up. Nineteen of them were reported by the reference member to have died, 16 from illness, and three unknown causes. Thirty-one participants left the study due to changes in household composition. The remaining 191 participants did not report any reasons for leaving the study.

### Baseline characteristics

Among the 6146 adults in the analysis, 3213 were female (52%) and 2933 males (48%), their mean age was 53 ± 15, and 62% of the participants were aged 50-74. More than half reported excellent self-perceived physical (54%) and mental (61%) health (Table 1). In previous studies, we showed that MAVIE participants were older and had higher socioeconomic and educational levels than the French population (Rojas Castro et al., 2021). The median percentage of time spent at home awake was 48% (Q1=36%, Q3=58%). Adults older than 50, unemployed, retirees, homemakers, and those who reported gardening, DIY, and domestic activities were those who spent the most time at home.

**Table 1.**
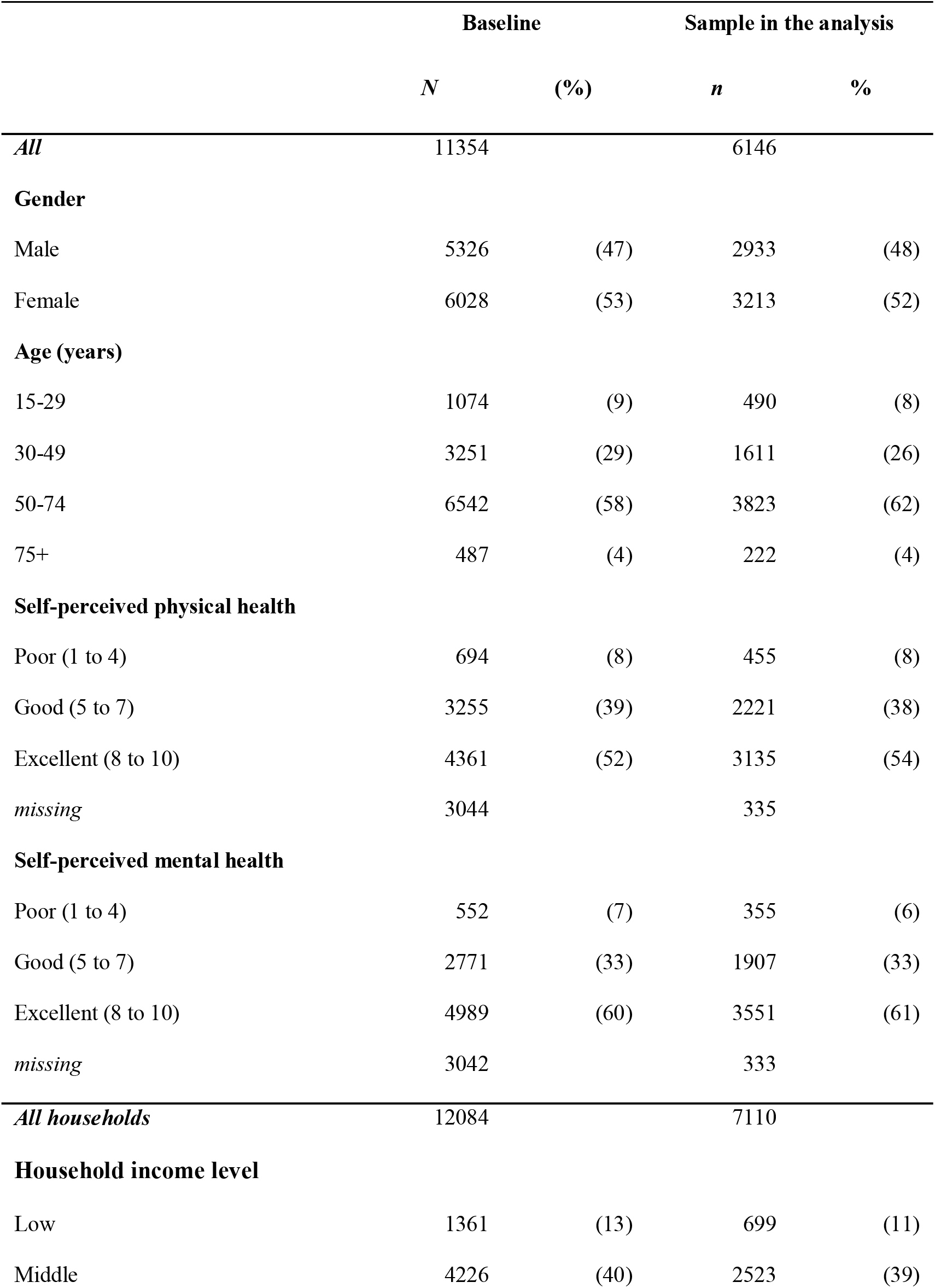
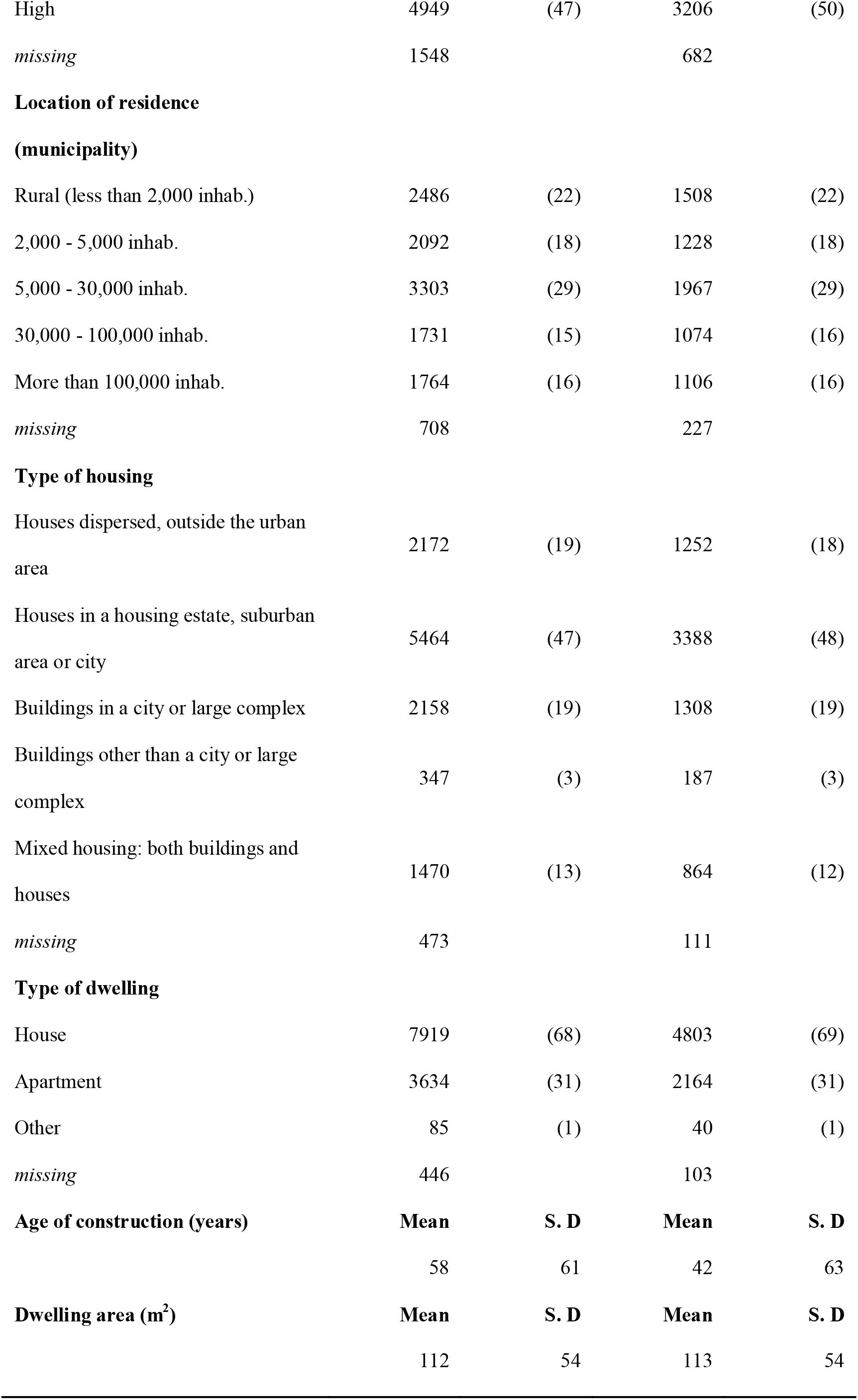
Comparison of the distributions of age, gender, self-perceived physical and mental health and general characteristics of the households between the baseline and analysed sample.

Most participants lived in a house (69%), mainly in cities or suburban areas, and 69% lived in municipalities of less than 30000 inhabitants. The average age of the construction was 42 ± 63 years, and the dwelling area was 113 ± 54 m^2^. Eight percent of the participants lived in constructions from before 1900. Most households had a personal outdoor space (70%) and a garage, box or garden hut (75%) with potentially dangerous objects, products and tools. More than half of the residences had stairs (58%).

The characteristics of the individuals and households of the study sample and the baseline sample were similar (Table 1). However, dropout was more frequent among participants aged 15 to 49 and among households with low-income levels. The dwellings of the analysis sample were also more recent than those of the baseline sample.

### Home Injuries

During follow-up, 738 (12%) participants reported at least one HI and 979 HI in total. Among these946 occurred while awake. Most of HIs were minor injuries, and only 28% required either hospitalization or emergency care. Falling was the main mechanism of HI (39%), domestic and gardening activities (42%), the most frequent activities, and the garden (25%) the most frequent location.

### Home injury risk factors

Among the 58-home environmental, behavioural and domestic activities factors considered (Table S1), the following were significantly associated with a higher risk of HI: storing household products out of their original packaging and using a stool to reach high places. No environmental characteristics of the dwelling were found significantly associated with a higher risk of HI.

After adjustment for confounding factors (Block 1, Table 2), DIY practices (occasional RR = 1.48; 95% CI, 1.12 - 1.94 or frequent RR= 1.75; 95% CI, 1.21 - 2.53), storing household products out of their original packaging (RR = 1.43; 95% CI, 1.07 - 1.92), and using a stool to reach high places (RR = 1.42; 95% CI, 1.10 - 1.83) were associated with a higher risk of HI in adults of all ages, with a small to moderate effect (Block 2, Table 2). Among the adjustment variables, a history of previous injury, high household income and living alone were also significantly associated with the risk of HI in all adults (Block 1, Table 2).

**Table 2.**
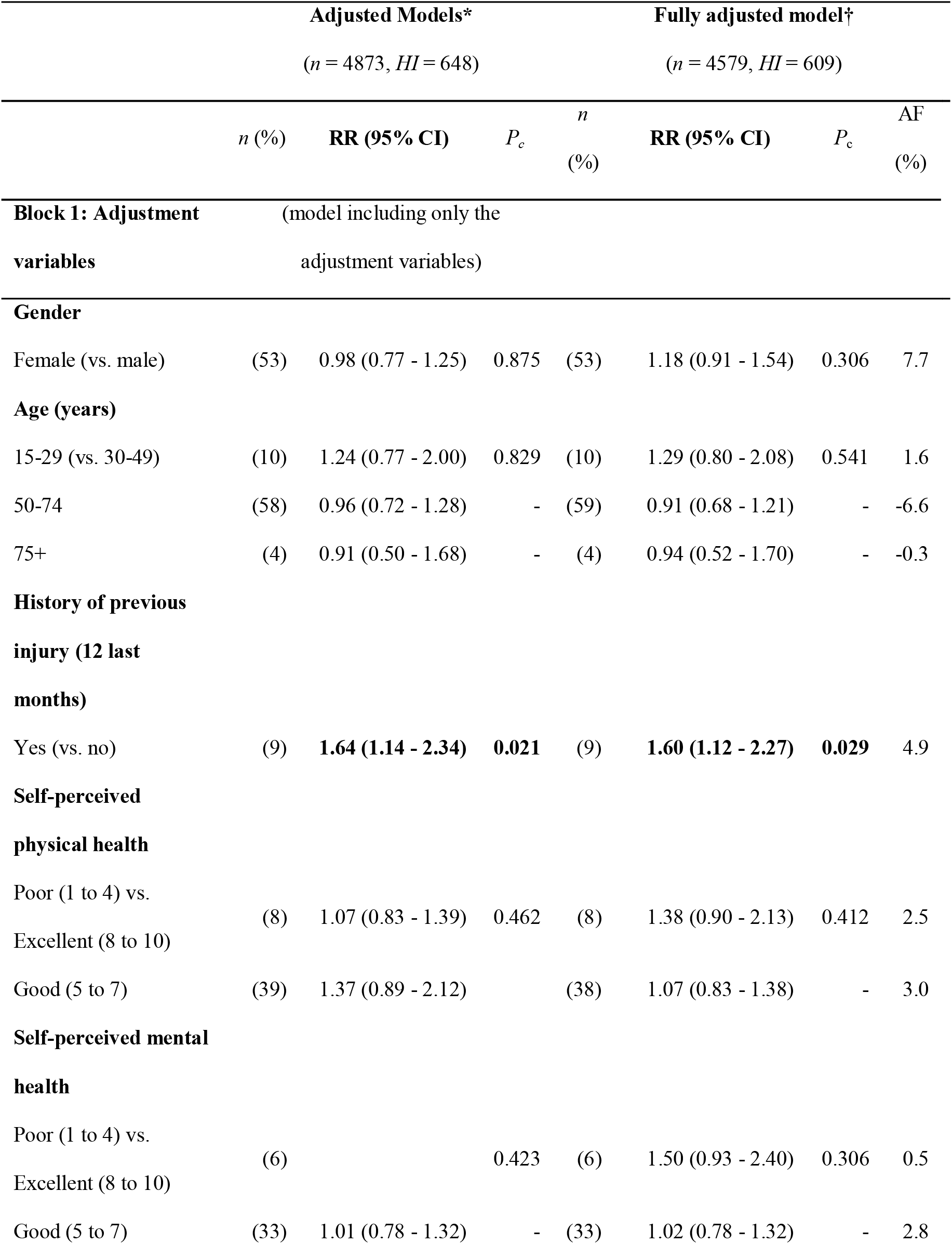
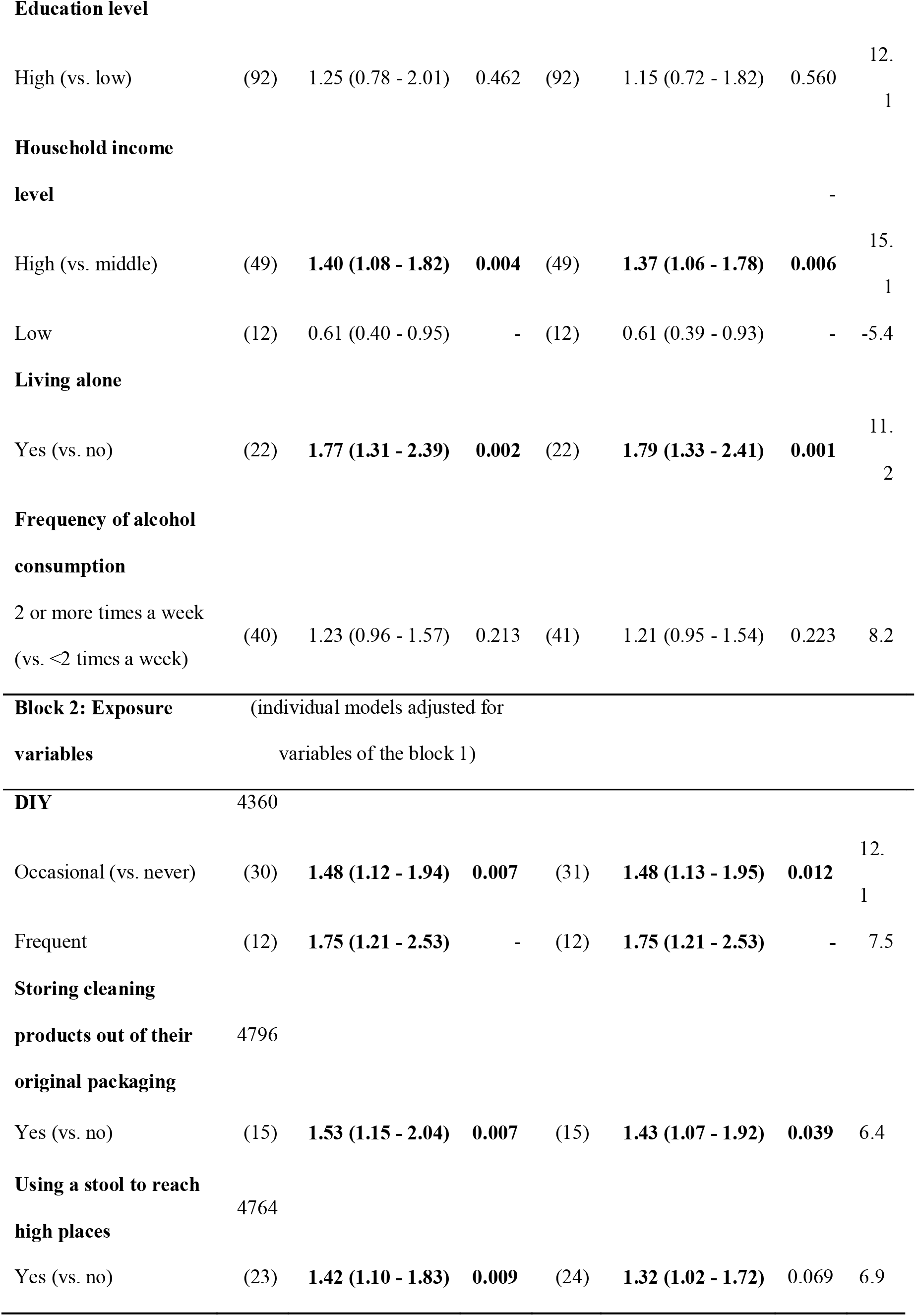
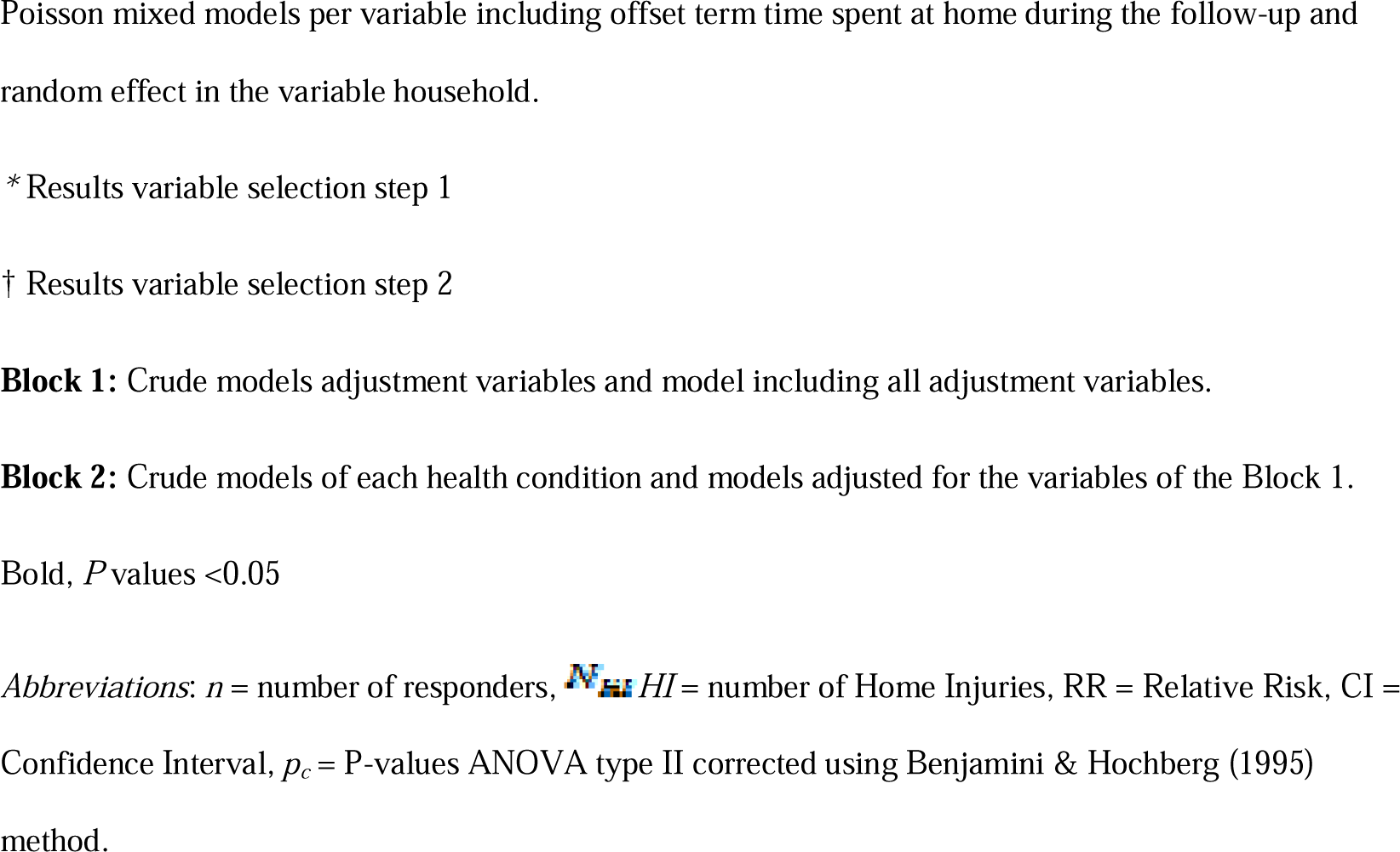
Home environmental, behavioural and domestic activities factors associated with the incidence of HI, relative to the time at risk at home, in adults of the MAVIE cohort (adjusted and fully adjusted models)

In the fully adjusted model (Table 2), DIY and storing household products outside their original packaging remained associated with HI risk with an AF of 12.1% and 6.4%, respectively. The risk contribution of using a stool to reach high places also remains significant (AF=6.9%), although with a weaker association (RR=1.32; 95% CI, 1.02-1.72).

### Age stratification

In adults under 50 years of age, we found that none of the factors assessed were significantly associated with the HI risk after adjustment. In adults aged 50 years of age and over, DIY, use of a stool to reach high places and the presence of stairs were significantly associated with HI (Table S2). DIY was the characteristic with the highest AF (11.1%). Among the adjustment variables, a history of previous injury, self-perceived poor mental and physical health, high household income and living alone were also significantly associated with the risk of HI in adults aged 50 years of age and over (Table S2).

## Discussion

Here, we present the first study to control for exposure times when assessing the association between home environmental, behavioural and domestic activity factors with the risk of non-fatal HIs.

We followed up 6146 cohort participants residing in French households, of whom 12% reported at least one HI over an average of 5.1 years. In adults of all ages, among the 58 variables evaluated, only the *storage of household products outside their original packaging* is associated with an increased risk of HI. In adults aged 50 and over, *DIY* and *stools used to reach high places* were associated with an increased risk of HI. In adults under 50 years of age, we found no association of exposures of interest with HI. A large and unprecedented number of factors of the physical environment were collected in our study. Despite this, we did not detect any association of these factors with the risk of HI. This result suggests that interventions targeting adults of all ages should not preferentially focus on modifying the physical environment. Instead, it suggests that behaviors play an important role in the risk of HI and in particular among adults aged 50 and over.

Changing the physical environment and mapping hazards in the homes of people with reduced mobility continue to be helpful to keep them safely active (Carnemolla & Bridge, 2015). The effectiveness of environmental modification interventions to reduce falls in older adults with reduced physical capacity is indeed well described (Gillespie et al., 2009, 2012).

In households where household products are usually stored out of their original packaging, the increase in the HI risk does not seem solely related to the increase in the number of poisonings. Similarly, among people aged 50 and over who reported using stools to reach high places in their homes, the proportion of HI was higher for falls but also for other mechanisms of injury. These factors may be ‘*markers’* of a lack of knowledge, risk underestimation or careless attitudes towards home safety. The high percentages of participants reporting these behaviours (15% *store household products out of their original packaging* and 22% *use stools to reach high places*) suggest that awareness campaigns on domestic risks is a relevant topic.

We confirmed the increased risk of HI associated with DIY practice in adults aged 50 and over (Ashby et al., 2007; Yiengprugsawan et al., 2012). In our study, the risk of HI was 63% higher among frequent DIYers and 40% higher among occasional DIYers, compared to people who did not report it. DIY was most common among people aged 50-69 years and retired people. Crushing and overexertion were the most frequent HI mechanisms (30.6% and 17.2%). The lack of experience and knowledge, underestimation of risks, overconfidence and cost of services are factors that can increase vulnerability to DIY injuries (Ashby et al., 2007; Verrier & Chevalier, 2007). Work at home could be another reason (Driscoll et al., 2003), but less probable in adults over-50s.

We detected a slight, non-significant association between gardening activities and the risk of HI. Consistent with an original study conducted in Switzerland (Schaudt et al., 2019), gardening-related HI were very frequent but most with minor consequences. The authors of this study highlight the incorrect use of gardening tools as a frequent cause of injuries (Schaudt et al., 2019). We found no association of HI with domestic activities such as cleaning and/or cooking.

Intervention focuses on DIY, gardening or of any other domestic activities should not discourage their practice. Leisure activities that keep people active have beneficial effects on health and wellbeing (Han & Patterson, 2007; Jiang & Xu, 2014), one example is the moderate practice of gardening (Thompson, 2018). Interventions may inform about existing risks, improve skills, promote risk self-assessment and self-management, such as the use of protective equipment.

A secondary result of this study, consistent with previous literature (Camilloni et al., 2011; Ferrante et al., 2014) was an increased risk of HI (86% higher) for people aged over-50s living alone. This higher risk may however be related with the involvement of older people in DIY.

An important number of aged people choose DIY to improve their general fitness, to feel satisfied and proud of a job well done, and to give meaning and pleasure to everyday tasks (Ashby et al., 2007). However, accumulated health problems and functional impairments increase the risk of many tasks, such as working at heights (Ashby et al., 2007).

Others, despite their state of health or lack of experience, may be forced to take risks because they live alone, do not have access to appropriate resources and services, or are afraid to use them for fear of financial or even physical abuse. (Ashby et al., 2007). The lack of maintenance would expose the person to other hazards. Accessible alternatives should be a priority when self-regulation is insufficient.

The lack of associations in adults under 50 years of age with any of the potential risk factors assessed in this study may be explained by the adjument on perceived health. The next step would be to investigate an integrated model that includes possible interactions between health, environmental and behavioural factors.

### Limitations and Strengths

Common limitations observed in volunteer-based cohorts and e-cohorts are low response rates, volunteer bias, loss of follow-up, and self-administered questionnaires, leading to selection biases and missing and selective answers. As with other e-cohorts (Nittas et al., 2021), we made continuous efforts to address representativeness, data reliability, under-reporting of the outcome, and loss to follow-up (Kristman et al., 2004; Rojas et al., 2021).

We mitigated problems related to self-reported information by using an online survey designed to minimize missing or inaccurate data, removing non-essential questions, improving user-friendliness, using intermittent saving options, and checking the consistency of answers to the different questions. Moreover, we used several strategies to reduce under-reporting of the outcome and loss of follow-up, such as quarterly email reminders and one-click event notifications. We estimated a follow-up rate of 86% (Rojas et al., 2021), which is higher than the 60-80% range considered acceptable by some authors for a valid study (Kristman et al., 2004).

Our sample is not representative of the French population. Young adults and people from low socioeconomic groups and low levels of educational attainment were underrepresented (Rojas et al., 2021). Dwellings with security problems are also under-represented. In a group of volunteers motivated to participate in a health study, socioeconomic and lifestyle differences between exposed and unexposed individuals would be less critical than in a representative sample of the general population (Nohr & Olsen, 2013; Richiardi et al., 2013; Rothman et al., 2013). Even if exposure heterogeneity was not captured, the greater homogeneity of the control factors might have increased the statistical power to detect main effects. Homogeneity may be desired when the multifactorial aspect is analysed.

We cannot rule out the existence of misclassification, especially for characteristics perceived as of minor importance. We think that the involvement of a reference person to communicate household information might increase the reliability of the data. The high follow-up rate also suggests that participants are motivated, increasing the reliability of the responses.

This study design does not allow us to disentangle the effects of risk associated with the activity itself and the risk of the tool or the product. We do not have information on the individual use of those products. We measured exposures at baseline; modifications in the environment, behaviours or the practice of activities were not assessed over time. Moreover, information about individual risk-taking and risk awareness was also not available. The sample size and the number of events prevented analyses in other population groups of interest, such as children and people with disabilities and different injury mechanisms.

## Conclusion

This study suggests that improving risk perception and self-regulation may be a key target for the development of effective interventions to reduce the number of HI. Interventions aimed at making DIY safer are likely to reduce HI. Awareness-raising campaigns on HI are still needed and enable them to manage them themselves.

## Supporting information

Supplemental Figure 1. Modelling steps

Supplemental Table 1. Crude models

Supplemental Table 1. Adjusted models, aged 50 or more

Supplementary material. Poisson mixed model with DHARMa R package (code and output)

## Data Availability

Personal health data underlying the study are protected by the French Data Protection Act and cannot be shared publicly. The large number of variables allows data to be indirectly identifiable, and making such data freely
available is prohibited.
Furthermore, an authorization from the CNIL, the French data protection authority, may be required to transfer the data, especially abroad. Data from this study can be obtained upon request from the
steering committee, as well
as from the corresponding author (Emmanuel Lagarde).

http://www.observatoire-mavie.com/contacter-equipe-MAVIE.aspx

