## Supplemental Figure 1. Modelling steps for "Effects of home environmental, behavioural and domestic activities on the risk of home injuries in French adults: Results from a prospective study"

### Modelling steps for the study of associations in relation to the physical environment, activities and behaviours and home injuries

**Step 0**

- Identify the main exposures of interest and confounding factors

$E = \{E_1, ..., E_p\}$ are the set of exposures of interest: the physical environment, activities and behaviours that can potentially be associated with home injuries.

$D = \{D_1, D_2, D_3, D_4, D_5\}$ are the confounding factors forced into the models: gender, age, history of previous injury, self-perceived physical and mental health status.

$C = \{C_1, ..., C_q\}$ are other confounding factors identified through *Directed Acyclic Graphs*.

**General model definition**

$$log(\mu_{ijk}) = log(t_{ijk}) + D_{ijk1}\alpha_1 + D_{ijk2}\alpha_2 + D_{ijk3}\alpha_3 + D_{ijk3}\alpha_3 + D_{ijk4}\alpha_4 + C_{ijk1}\gamma_1 + ... + C_{ijkq}\gamma_q + E_{ijk1}\beta_1 + E_{ijkq}\beta_q + b_k + b_{jk}, \quad (1)$$

where $\mu_{ijk}$ is the expectation of the home injury incidence of multiple measurements of each of the households where the same individual has lived $i$, the individual $j$, belonging to the same household $k$, $b_k$ the random intercept of the household $k$, $b_{jk}$ the nested random intercept of individuals in the household, $log(t_{jk})$ is the term offset.

- Estimation of models for each factor: gender, age, history of previous injury, self-perceived physical and mental health status
- Estimation of $p$ models containing a single exposure of interest without adjustment
- Estimation of $q$ models containing a single factor without adjustment

**Step 1**

- Estimation of $q$ models containing a single confounder adjusted for gender, age, history of previous injury, self-perceived physical and mental health status

$$log(\mu) = log(t) + D_1 + D_2 + D_3 + D_4 + D_5 + C_1 + b, \quad (2)$$

$$\vdots$$

$$log(\mu) = log(t) + D_1 + D_2 + D_3 + D_4 + D_5 + C_q + b.$$

- Selection of significant potential $q^*$ confounders after Benjamini and Hochberg (BH) correction ($P < 0.1$) adjusted for gender, age and history of previous injury.
- Estimation of a model containing $q^*$ retained confounders adjusted for gender, age and history of previous injury, self-perceived physical and mental health status:

$$log(\mu) = log(t) + D_1 + D_2 + D_3 + D_4 + D_5 + C_1 + ... + C_{q^*}. \quad (3)$$

- Parameter Estimation, Confidence Intervals (CI), $P$-value

- Estimation of p models containing a single exposure of interest adjusted for gender, age and history of previous injury

$$log(\mu) = log(t) + D_1 + D_2 + D_3 + D_4 + D_5 + E_1 + b, \quad (4)$$

$$\vdots$$

$$log(\mu) = log(t) + D_1 + D_2 + D_3 + D_4 + D_5 + E_p + b.$$

- Selection of $p^*$ significant potential confounders after BH correction ($P < 0.1$) adjusted for gender, age, history of previous injury, self-perceived physical and mental health status.

**Step 2**

- Estimation of $p^*$ models containing 1 single exposure adjusted for gender, age, history of previous injury, and the selected $q^*$ confounders:

$$log(\mu) = log(t) + D_1 + D_2 + D_3 + D_4 + D_5 + C_1 + ... + C_{q*} + E_1 + b, \tag{5}$$

$$\vdots$$

$$log(\mu) = log(t) + D_1 + D_2 + D_3 + D_4 + D_5 + C_1 + ... + C_{q*} + E_{p^*} + b.$$

- Selection of $p^{**}$ significant BH-corrected exposures ($P < 0.1$) adjusted for gender, age, history of previous injury, self-perceived physical and mental health status of the $q*$ confounders retained.
- Parameter estimates, CIs, adjusted $P$-value BH

**Step 3**

- Estimation of a model containing $p^{**}$ exposures adjusted for gender, age, history of previous injury and the selected $q^*$ confounders:

$$log(\mu) = log(t) + D_1 + D_2 + D_3 + D_4 + D_5 + C_1 + ... + C_{q*} + E_1 + ... + E_{p^{**}} + b. \tag{6}$$

- Parameter estimate, CI, $P$-value$< 0.1$, Attributable fraction

In all models, in order to simplify the notations, when selecting for example the $p^*$ factors from $E1, ...Ep$, the exposures are reordered in such a way that the selected $p^*$ exposures are placed first: $E1, ...E_{p^*}$.
