## Supplemental Table 1. Crude models for "Effects of home environmental, behavioural and domestic activities on the risk of home injuries in French adults: Results from a prospective study"

| | | Crude Models ($n$ = 6146, $HI$ = 946) | |
|---|---|---|---|
| | $n$ (%) | RR (95% CI) | $p_c$ |
| **Block 1: Adjustment variables** | | | |
| **Gender** | 6146 | | |
| Female (vs. male) | (52) | 1.02 (0.84 - 1.23) | 0.852 |
| **Age (years)** | 6146 | | |
| 15-29 (vs. 30-49) | (8) | 1.04 (0.70 - 1.55) | 0.706 |
| 50-74 | (62) | 0.88 (0.70 - 1.10) | - |
| 75+ | (4) | 0.93 (0.55 - 1.54) | - |
| **History of previous injury (12 last months)** | 5301 | | |
| Yes (vs. No) | (9) | 1.95 (1.41 - 2.70) | **0.004** |
| **Self-perceived physical health** | 5811 | | |
| Poor (1 to 4) (vs. 8 - 10) | (8) | 1.61 (1.14 - 2.25) | 0.110 |
| Good (5 to 7) | (38) | 1.11 (0.91 - 1.37) | - |
| **Self-perceived mental health** | 5813 | | |
| Poor (1 to 4) (vs. 8 - 10) | (6) | 1.57 (1.08 - 2.29) | 0.211 |
| Good (5 to 7) | (33) | 1.06 (0.86 - 1.31) | - |
| **Level of educational attainment by age** | 6132 | | |
| High (vs. Low) | (92) | 1.54 (1.03 - 2.31) | 0.148 |
| **Household income level** | 5510 | | |
| High (vs. Middle) | (45) | 1.24 (1.00 - 1.53) | 0.110 |
| Low | (10) | 0.79 (0.55 - 1.14) | - |
| **Living alone** | 6098 | | |
| Yes (vs. No) | (20) | 1.36 (1.09 - 1.71) | 0.066 |
| **Frequency of alcohol consumption** | 5918 | | |
| 2 or more times a week (vs. <2 times a week) | (42) | 1.17 (0.96 - 1.42) | 0.314 |
| **Employment status** | 6086 | | |
| Unemployed, Homemakers or Retirees (vs. Students, Employees) | (51) | 0.82 (0.67 - 1.00) | 0.179 |
| **Block 2: Exposures of interest** | | (one model per variable) | |
| ***Household characteristics*** | | | |
| **Year construction** | 5383 | | |
| <1900 (vs. >=2000) | (8) | 1.39 (0.96 - 2.02) | 0.285 |
| 1900-1949 | (11) | 1.36 (0.96 - 1.93) | - |
| 1950-1979 | (27) | 1.07 (0.80 - 1.42) | - |
| 1980-1999 | (23) | 0.95 (0.70 - 1.28) | - |
| **House area** | 6008 | | |
| <50 vs. (90-129) | (6) | 1.38 (0.91 - 2.11) | 0.699 |
| 50-89 | (26) | 1.06 (0.82 - 1.36) | - |
| 130-169 | (17) | 1.06 (0.81 - 1.39) | - |
| >=170 | (12) | 1.16 (0.86 - 1.57) | - |
| **Animals** | 6033 | | |

| | | | |
|---|---|---|---|
| Yes (vs. No) | (43) | 1.14 (0.94 - 1.39) | 0.396 |
| **Heating** | 5933 | | |
| Individuel (vs. Collective) | (85) | 1.17 (0.85 - 1.61) | 0.560 |
| **Fireplace** | 5883 | | |
| Yes (vs. No) | (15) | 0.87 (0.67 - 1.12) | 0.527 |
| **Date maintenance heating** | 4823 | | |
| More than 1 year (vs. Less than 1 year) | (11) | 0.98 (0.72 - 1.33) | 0.648 |
| More then 5 years | (5) | 1.26 (0.85 - 1.86) | - |
| **Auxiliary heating** | 5885 | | |
| Yes, regularly (vs. No, never) | (5) | 1.13 (0.73 - 1.75) | 0.779 |
| Yes, sometimes | (22) | 1.08 (0.86 - 1.35) | - |
| **Date electrical renovation** | 5012 | | |
| More than 40 years (vs. Less than 40 years) | (3) | 1.30 (0.75 - 2.25) | 0.560 |
| **Wires woven in the electrical installation** | 4431 | | |
| Yes (vs. No) | (2) | 0.86 (0.49 - 1.52) | 0.699 |
| **Adapted logement (disabled people)** | 5943 | | |
| Yes (vs. No, it is not useful) | (6) | 0.89 (0.60 - 1.32) | 0.852 |
| No, but it would be useful | (7) | 1.01 (0.70 - 1.46) | - |
| **Absence of smoke alarms** | 5801 | | |
| Yes (vs. No) | (20) | 0.91 (0.71 - 1.16) | 0.619 |
| **Balcony** | 4146 | | |
| Yes (vs. No) | (30) | 0.76 (0.61 - 0.94) | 0.096 |
| **Weapon** | 5961 | | |
| Yes (vs. No) | (18) | 1.11 (0.88 - 1.41) | 0.571 |
| **Parquet on the kitchen floor** | 5856 | | |
| Yes (vs. No) | (5) | 1.18 (0.76 - 1.82) | 0.619 |
| **Linoleum on the kitchen floor** | 5854 | | |
| Yes (vs. No) | (11) | 1.09 (0.80 - 1.48) | 0.699 |
| **Tile on the kitchen floor** | 5923 | | |
| Yes (vs. No) | (82) | 0.93 (0.71 - 1.22) | 0.699 |
| **Tapis on the room** | 6142 | | |
| Yes (vs. No) | (34) | 1.16 (0.95 - 1.42) | 0.110 |
| **Cable phone in the room** | 6142 | | |
| Yes (vs. No) | (47) | 1.26 (1.04 - 1.53) | 0.370 |
| **Not easy access to the lights from the bed** | 5726 | | |
| Yes (vs. No) | (6) | 0.86 (0.55 - 1.36) | 0.686 |
| **Low-light lamp in the room** | 5875 | | |
| Yes (vs. No) | (5) | 1.40 (0.95 - 2.08) | 0.283 |
| **Tiling on the living room or room floor** | 5870 | | |
| Yes (vs. No) | (61) | 1.08 (0.88 - 1.33) | 0.619 |
| **Linoleum on the living room or room floor** | 5826 | | |

| | | | |
|---|---|---|---|
| Yes (vs. No) | (21) | 0.93 (0.73 - 1.18) | 0.687 |
| **Carpet on the living room or room floor** | 5795 | | |
| Yes (vs. No) | (16) | 1.15 (0.90 - 1.48) | 0.522 |
| **Parquet on the living room or room floor** | 5889 | | |
| Yes (vs. No) | (67) | 1.20 (0.97 - 1.49) | 0.283 |
| **Bathtub** | 5927 | | |
| Yes (vs. No) | (68) | 1.10 (0.89 - 1.36) | 0.571 |
| **Shower** | 5930 | | |
| Yes (vs. No) | (67) | 0.91 (0.74 - 1.12) | 0.560 |
| **Linoleum on the bathroom floor** | 5864 | | |
| Yes (vs. No) | (15) | 1.14 (0.88 - 1.49) | 0.560 |
| **Tiles on the bathroom floor** | 5937 | | |
| Yes (vs. No) | (79) | 0.88 (0.69 - 1.12) | 0.539 |
| **Attic** | 5910 | | |
| Yes (vs. No) | (33) | 1.13 (0.92 - 1.38) | 0.497 |
| **Cellar** | 5862 | | |
| Yes (vs. No) | (44) | 1.18 (0.97 - 1.43) | 0.285 |
| **Stairs** | 5953 | | |
| Yes (vs. No) | (58) | 1.32 (1.08 - 1.62) | 0.066 |
| **Ramped stairs** | 123 | | |
| All of them (vs. None of them) | (56) | 1.02 (0.70 - 1.49) | 0.699 |
| Some of them | (10) | 1.23 (0.76 - 2.01) | - |
| **No lighting stairs** | 138 | | |
| None of them (vs. All of them) | (2) | 1.07 (0.45 - 2.51) | 0.571 |
| Some of them | (1) | 1.94 (0.77 - 4.91) | - |
| **No antislip stairs** | 149 | | |
| None of them (vs. All of them) | (65) | 1.72 (1.06 - 2.78) | 0.179 |
| Some of them | (5) | 1.20 (0.59 - 2.46) | - |
| **Garage, box or hut garden** | 5097 | | |
| Yes (vs. No) | (75) | 0.85 (0.62 - 1.16) | 0.539 |
| **Sharp objects** | 2419 | | |
| Yes (vs. No) | (71) | 0.70 (0.52 - 0.96) | 0.127 |
| **Toxic dangers** | 2028 | | |
| Yes (vs. No) | (55) | 0.90 (0.70 - 1.17) | 0.613 |
| **Heavy dangers** | 2258 | | |
| Yes (vs. No) | (61) | 1.08 (0.82 - 1.41) | 0.699 |
| **Irritating dangers** | 1789 | | |
| Yes (vs. No) | (46) | 0.85 (0.66 - 1.09) | 0.446 |
| **Flammable hazards** | 2122 | | |
| Yes (vs. No) | (59) | 0.95 (0.73 - 1.25) | 0.779 |
| **Explosive hazards** | 1058 | | |
| Yes (vs. No) | (15) | 1.05 (0.79 - 1.40) | 0.779 |
| **Corrosive hazards** | 1591 | | |
| Yes (vs. No) | (38) | 0.88 (0.69 - 1.13) | 0.539 |

| | | | |
|---|---|---|---|
| **Ladders** | 2236 | | |
| Yes (vs. No) | (64) | 0.74 (0.56 - 0.97) | 0.138 |
| **Electric tools** | 2532 | | |
| Yes (vs. No) | (74) | 0.65 (0.47 - 0.88) | 0.066 |
| **Manuals tools** | 2750 | | |
| Yes (vs. No) | (80) | 0.67 (0.47 - 0.94) | 0.110 |
| **Outdoor space (N=4,646)** | 5931 | | |
| Yes, for personal use (vs. No) | (70) | 1.30 (0.99 - 1.73) | 0.388 |
| Yes, as a communal area (vs. No) | (9) | 1.32 (0.88 - 1.98) | - |
| **Lawn mower** | 3207 | | |
| Yes (vs. No) | (64) | 1.15 (0.89 - 1.48) | 0.539 |
| **Barbecue** | 3177 | | |
| Yes (vs. No) | (59) | 1.11 (0.88 - 1.41) | 0.571 |
| **Uneven ground** | 3207 | | |
| Yes (vs. No) | (16) | 1.22 (0.94 - 1.59) | 0.344 |
| **Water source (including pools)** | 3451 | | |
| Yes (vs. No) | (68) | 0.87 (0.69 - 1.10) | 0.497 |
| ***Individual activities and behaviors*** | | | |
| **DIY** | 6627 | | |
| Occasional (vs. Never) | (29) | 1.30 (1.05 - 1.62) | 0.111 |
| Frequent | (11) | 1.41 (1.05 - 1.88) | - |
| **Gardening** | 5662 | | |
| Occasional (vs. Never) | (34) | 1.26 (1.02 - 1.57) | 0.151 |
| Frequent | (13) | 1.35 (1.03 - 1.79) | - |
| **Domestic activities** | 5550 | | |
| Occasional (vs. Never) | (53) | 1.45 (1.12 - 1.88) | 0.112 |
| Frequent | (19) | 1.36 (1.00 - 1.86) | - |
| **Decanting cleaning products from the original packaging** | 5951 | | |
| Yes (vs. No) | (16) | **1.53 (1.20 - 1.95)** | **0.014** |
| **Using a chair to reach high places** | 5914 | | |
| Yes (vs. No) | (38) | 1.32 (1.09 - 1.60) | 0.066 |
| **Using a ladder to reach high places** | 5927 | | |
| Yes (vs. No) | (67) | 0.97 (0.79 -1.20) | 0.852 |
| **Using a small ladder to reach high places** | 5917 | | |
| Yes (vs. No) | (43) | 1.18 (0.97 - 1.43) | 0.283 |
| **Using a stool to reach high places** | 5891 | | |
| Yes (vs. No) | (23) | **1.50 (1.22 - 1.86)** | **0.006** |
