## Supplemental Table 1. Adjusted models, aged 50 or more for "Effects of home environmental, behavioural and domestic activities on the risk of home injuries in French adults: Results from a prospective study"

**Supplementary material 2** Home environmental, behavioural and domestic activities factors associated with the incidence of HI, relative to the time at risk at home, in adults aged 50 or more in the MAVIE cohort

| | Adjusted Models* (*n* = 3024, *HI* = 465) | | | Fully adjusted model† (n = 2801, *HI* = 434) | | | |
|---|---|---|---|---|---|---|---|
| | *n* (%) | **RR (95% CI)** | $P_c$ | *n* (%) | **RR (95 % CI)** | *P* | AF (%) |
| **Block 1: Adjustment variables** | | (model including only the adjustment variables) | | | | | |
| **Gender** | | | | | | | |
| Female (vs. male) | (54) | 0.88 (0.68 - 1.15) | 0.546 | (53) | 1.05 (0.77 - 1.43) | 0.764 | 2.0 |
| **Age (years)** | | | | | | | |
| 75+ (vs. 50-74) | (6) | 0.90 (0.53 -1.54) | 0.703 | (6) | 1.06 (0.62 - 1.81) | 0.832 | 0.4 |
| **History of previous injury (12 last months)** | | | | | | | |
| Yes (vs. no) | (8) | 1.86 (1.26 - 2.76) | 0.006 | (8) | 1.73 (1.16 - 2.58) | 0.007 | 5.6 |
| **Self-perceived physical health** | | | | | | | |
| Poor (1 to 4) vs. Excellent (8 to 10) | (8) | 1.31 (0.80 - 2.15) | 0.551 | (8) | 1.33 (0.80 - 2.22) | 0.496 | 2.6 |
| Good (5 to 7) | (39) | 0.95 (0.71 - 1.27) | - | (39) | 1.00 (0.74 - 1.34) | - | -0.2 |
| **Self-perceived mental health** | | | | | | | |
| Poor (1 to 4) vs. Excellent (8 to 10) | (5) | 1.31 (0.80 - 2.15) | 0.057 | (5) | 2.11 (1.21 - 3.67) | 0.026 | 4.5 |
| Good (5 to 7) | (29) | 0.95 (0.71 - 1.27) | - | (30) | 1.01 (0.74 - 1.38) | - | 0.4 |
| **Level of educational attainment by age** | | | | | | | |
| High (vs. low) | (93) | 1.22 (0.69 - 2.14) | 0.551 | (93) | 1.16 (0.65 - 2.08) | 0.619 | 13.2 |
| **Household income level** | | | | | | | |
| High (vs. middle) | (53) | 1.62 (1.20 - 2.18) | 0.001 | (53) | 1.53 (1.13 - 2.08) | 0.003 | 21.5 |
| Low | (9) | 0.60 (0.34 - 1.04) | - | (9) | 0.66 (0.38 - 1.16) | - | -3.1 |
| **Living alone** | | | | | | | |
| Yes (vs. no) | (23) | 1.98 (1.42 - 2.76) | <0.001 | (23) | 2.04 (1.44 - 2.89) | <0.001 | 13.7 |
| **Frequency of alcohol consumption** | | | | | | | |
| 2 or more times a week (vs. <2 times a week) | (50) | 1.34 (1.03 -1.75) | 0.057 | (50) | 1.32 (1.00 - 1.73) | 0.050 | 13.9 |
| **Bloc 2: Exposure variables** | | (individual models adjusted for variables of the block 1) | | | | | |
| **DIY** | 2906 | | | | | | |
| Occasional (vs. Never) | (34) | 1.42 (1.03 - 1.95) | 0.089 | (35) | 1.40 (1.01 - 1.94) | 0.043 | 11.1 |
| Frequent | (16) | 1.73 (1.16 - 2.56) | - | (16) | 1.63 (1.08 - 2.44) | - | 8.3 |
| **Domestic activities** | 2881 | | | | | | |
| Occasional (vs. Never) | (51) | 1.20 (0.89 - 1.62) | 0.488 | - | - | - | - |

| | | | | | | | |
|---|---|---|---|---|---|---|---|
| Frequent | (21) | 1.28 (0.83 - 1.96) | - | - | - | - | - |
| **Storing cleaning products out of their original packaging** | 2988 | | | | | | |
| Yes (vs. No) | (16) | 1.43 (1.03 - 1.99) | 0.089 | (16) | 1.32 (0.94 - 1.86) | 0.112 | 5.1 |
| **Using a stool to reach high places** | 2978 | | | | | | |
| Yes (vs. no) | (22) | 1.52 (1.13 - 2.04) | 0.052 | (23) | 1.42 (1.04 - 1.94) | 0.028 | 8.6 |
| **Using a chair to reach high places** | 2982 | | | | | | |
| Yes (vs. no) | (28) | 1.32 (0.99 - 1.75) | 0.096 | (29) | 1.16 (0.86 - 1.57) | 0.329 | 4.4 |
| **Parquet on the kitchen floor** | 2981 | | | | | | |
| Yes (vs. no) | (69) | 1.20 (0.89 - 1.62) | 0.249 | - | - | - | - |
| **Balcony** | 2132 | | | | | | |
| Yes (vs. no) | (30) | 0.78 (0.58 - 1.05) | 0.138 | - | - | - | - |
| **Cellar** | 2952 | | | | | | |
| Yes (vs. no) | (47) | 1.32 (1.01 - 1.71) | 0.089 | (48) | 1.30 (1.00 - 1.71) | 0.052 | 12.8 |
| **Stairs** | 2995 | | | | | | |
| Yes (vs. no) | (63) | 1.34 (1.01 - 1.79) | 0.089 | (64) | 1.31 (0.97 - 1.78) | 0.081 | 16.6 |
| **No lighting stairs** | 2615 | | | | | | |
| None of them (vs. All of them) | (30) | 3.19 (0.91 - 11.17) | 0.191 | - | - | - | - |
| Some of them | (41) | 1.59 (0.47 - 5.41) | - | - | - | - | - |

Poisson mixed models per variable including offset term time spent at home during the follow-up and random effect in the variable household.

* Results variable selection step 1

† Results variable selection step 2

**Block 1:** Crude models adjustment variables and model including all adjustment variables.

**Block 2:** Crude models of each health conditions and models adjusted for the variables of the Block 1.

Bold, *P* values <0.05

*Abbreviations*: *n* = number of responders, *HI* = number of Home Injuries, RR = Relative Risk, CI = Confidence Interval, $P_c$ = P-values ANOVA type II corrected using Benjamini & Hochberg (1995) method.
