## Supplementary material. Poisson mixed model with DHARMa R package (code and output) for "Effects of home environmental, behavioural and domestic activities on the risk of home injuries in French adults: Results from a prospective study"

### Supplementary Material 3

July 17, 2022

**Model mixte poisson and validation with DHARMA package**

```
#Algorithm TMB ------------------------------------------------------------
library(glmmTMB)

# Model using glmmTMB algorithm: mix model in household and participant

modelmixt_poisson <-
glmmTMB(n_injuries_awake ~
offset(log(y_maison_not_sleep)) + #time at home awake
(1|DEM_STRCLE)+ #Id foyer
(1|DEM_STRCLE:IND_CLE)+ #ID participant
IND_SEXE + #gender
age_cat + #age categories
hospit_acc + #history of previous injury
etat_sante_mental_cat + #self-reported mental health
etat_sante_physique_cat + #self-reported physical health
revenu_annuel_deciles + #household income level
alone + #living alone
diplome_eleve_cat2 + #education level
freq_alcool_cat +#frequency of alcohol consummation
tps_activite6_cat + # DIY
produits_menagers_transvaser + #storing cleaning products out of their ori
logement_lieux_eleves_tabouret, #using a stool to reach high places
data_model_final,
family = poisson(link = "log"), ziformula = ~ 0)

summary(modelmixt_poisson)
```

```
# Models validation: residuals diagnostic for mixed regression models

library(DHARMa)

# simulation of scaled residuals for GLMM scaled 0 to 1
# interpreted as linear regression residuals

simulationOutput_poisson <-
DHARMa::simulateResiduals(fittedModel = modelmixt_poisson)

#plotting the scaled residuals

diagnostic_plot_poisson <-
plot(simulationOutput_poisson, quantreg = FALSE)

#Goodness-of-fit test on scaled residuals------------------------------

#plotting the scaled residuals (visual aid to detect deviations):

#qq-plot to detect overall deviation from the expected distribution and
# Kolmogorov-Smirnov test.

#plot of the residuals against the predicted value

DHARMa::testResiduals(simulationOutput_poisson)

#Test uniformity: test if the overall distribution conform to expectation

#Test of zero inflation: test if there are more zeros in the data
# than expected from the simulations.

#Test of dispertion: test if the simulated dispersion is equal
# to the observed dispersion

#Test of outliers: test if there are more simulation outliers
#than expected

#Plots ------------------------------------------------------------------
```

```r
par(mfrow = c(1, 3))
#Test of zeroinflation
DHARMa::testZeroInflation(simulationOutput_poisson)

#Test of dispertion
DHARMa::testDispersion(simulationOutput_poisson)

#Test of outliers
DHARMa::testOutliers(simulationOutput_poisson)
par(mfrow = c(1, 1))
```

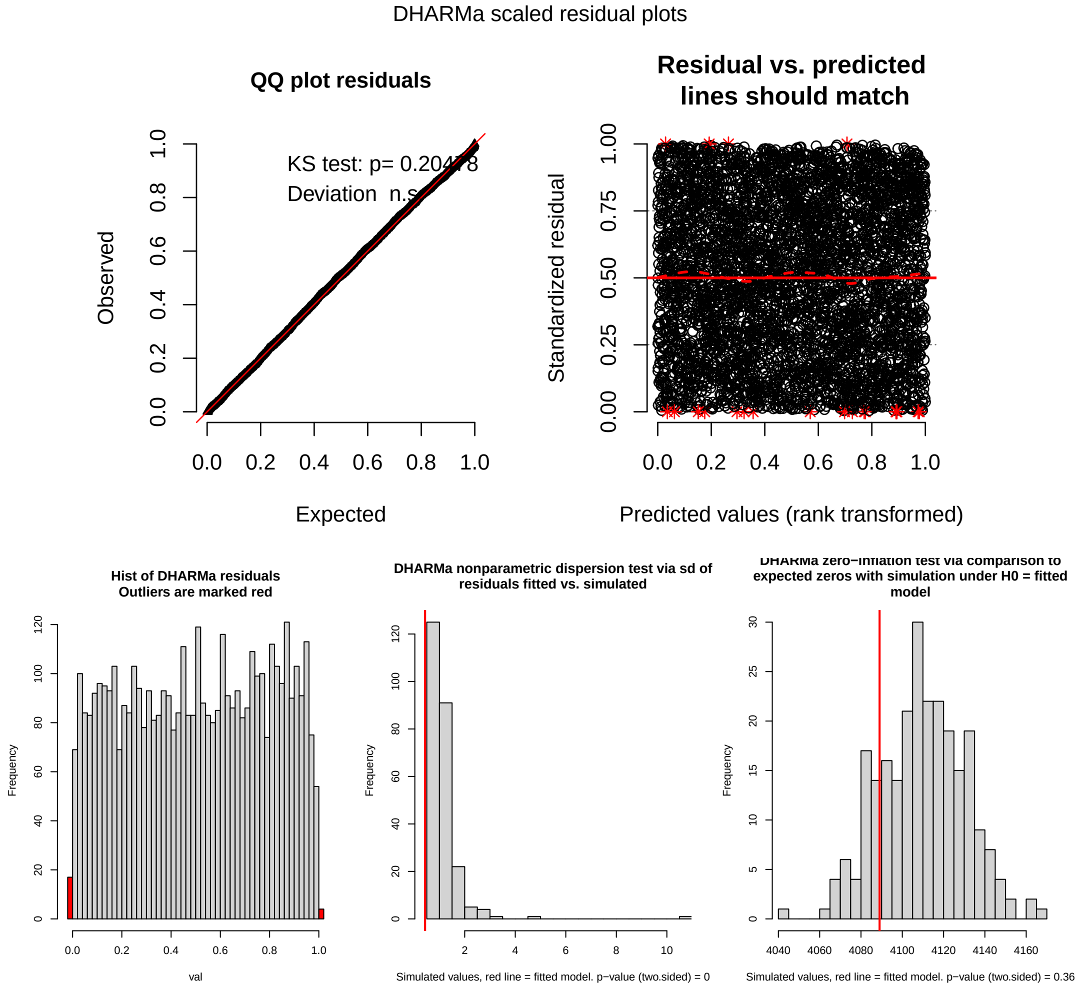


Figure 1: **Diagnostic plots of the Poisson model with mixed effects (complete model).** Uniformity test: Kolmogorov-Smirnov (valeur $p = 0.343$), test zero inflation (valeur $p = 0.360$), test dispersion (valeur $p < 0.001$), test outliers (valeur $p = 0.999$).

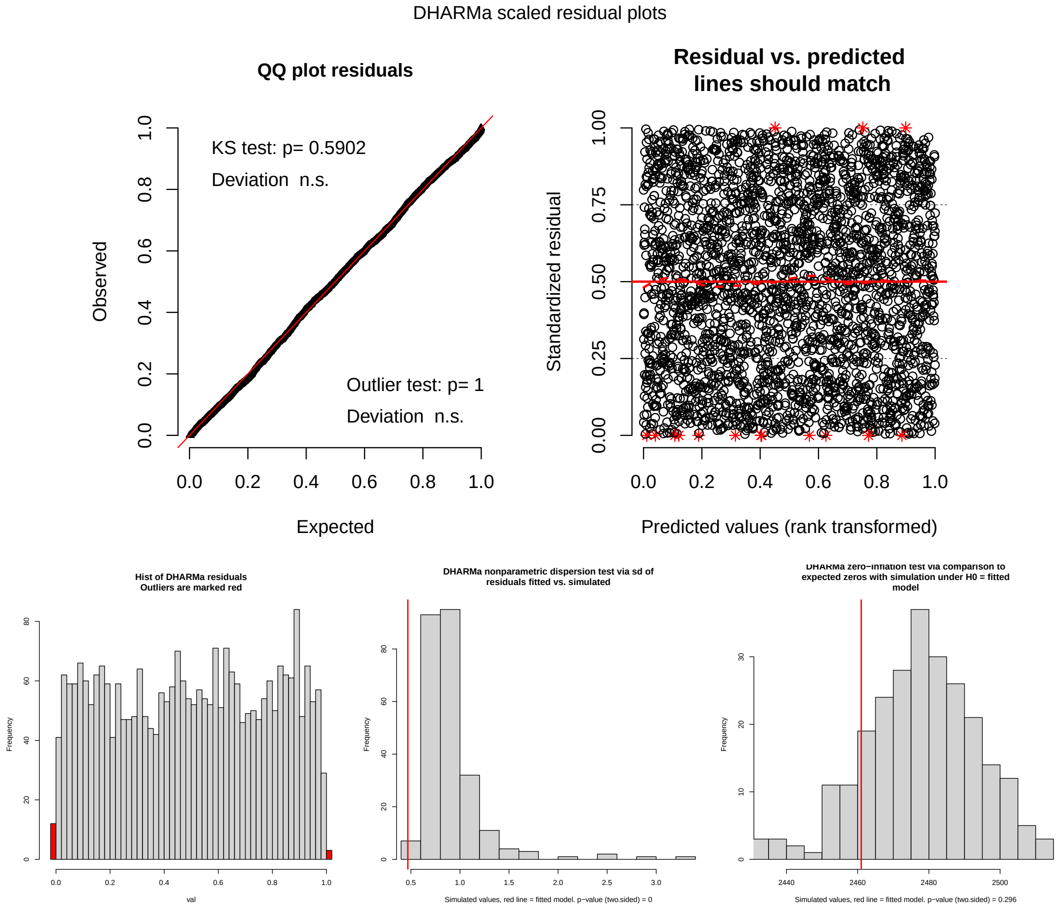


Figure 2: **Diagnostic plots of the Poisson model with mixed effects 50 years and more (complete model)**. Uniformity test: Kolmogorov-Smirnov (valeur $p$ = 0.590), test zero inflation (valeur $p$ = 0.296), test dispersion (valeur $p$ <0.001), test outliers (valeur $p$ = 0.999).
